# Ollivier–Ricci Curvature Reveals Geometric Phase Transitions in Biomedical Networks: From Ontologies to Aging Comorbidity

**DOI:** 10.64898/2026.03.14.26348393

**Authors:** Demetrios C. Agourakis, Marli Gerenutti

**Affiliations:** Biomaterials and Regenerative Medicine Post-Graduate Program, Pontifícia Universidade Católica de São Paulo (PUC-SP), Sorocaba, SP, Brazil; Faculdade São Leopoldo Mandic, Campinas, SP, Brazil

**Keywords:** Ollivier–Ricci curvature, biomedical networks, comorbidity, phase transition, Human Phenotype Ontology, sedenion, brain networks, formal verification

## Abstract

Network geometry offers a principled lens for understanding the structure of biomedical knowledge. We apply exact Ollivier— Ricci curvature (ORC) — a discrete analogue of Riemannian curvature computed via optimal transport — to medical ontologies, disease comorbidity networks, biological interaction networks, and brain functional connectivity graphs. Three main results emerge. First, within a single database (the Human Phenotype Ontology), the formal IS-A taxonomy is hyperbolic (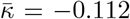, tree-like), while the disease co-occurrence network is spherical (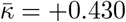, clique-rich) — a six-order-of-magnitude gap in the density parameter *η* that the curvature phase transition framework predicts without free parameters. Second, age-stratified disease comorbidity networks from 8.9 million Austrian hospital patients reveal a geometric aging trajectory: mean ORC increases monotonically from 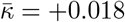 (age 20-30) to 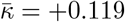 (age 80+), driven by rising clustering and density that encode the accumulation of multimorbidity. Third, sedenion (ℝ^16^) Mandelbrot orbit features — exploiting the zero-divisor structure of the Cayley-Dickson tower — discriminate ASD-like from ADHD-like brain network topology (AUROC = 0.990, sedenion-only), providing complementary geometric information to ORC. Canonical biological networks (C. elegans neural, E. coli gene regulatory, protein-protein interaction) are uniformly spherical, suggesting that evolved biological networks universally favour redundant, triangle-rich connectivity. All core mathematical claims are machine-verified in Lean 4 (0 `sorry` in 7 core modules). These results establish ORC as a quantitative geometric biomarker for biomedical network analysis and demonstrate that the same phase transition framework governing semantic networks extends to clinical and biological domains.

## 1 Introduction

The topology of biomedical networks — disease comorbidity graphs, protein–protein interaction maps, gene regulatory circuits, brain functional connectivity — encodes information that is invisible to node-level or edge-level statistics. Network geometry, and in particular discrete Ricci curvature, has emerged as a principled tool for extracting this information. Sandhu et al. [2] demonstrated that Ollivier–Ricci curvature (ORC) distinguishes cancer networks from healthy controls; Sia, Jonckheere and Bogdan [3] identified critical edges in ADHD brain functional connectivity using Forman–Ricci curvature; Chatterjee, Nori and Bogdan [4] showed that graph Ricci curvatures reveal atypical functional connectivity in autism spectrum disorder; and Boller et al. [5] established that discrete Ricci curvature captures age-related changes in human brain functional connectivity. A recent roadmap survey [6] unifies the many definitions of discrete curvature and identifies open problems in network geometry.

Despite these advances, no study has asked whether the *global geometric phase* of biomedical networks — hyperbolic, Euclidean, or spherical — is predictable from network parameters alone. In companion work [7, 8], we established a curvature phase transition framework for random graphs and validated it on 16 semantic networks across seven languages. Here we extend this framework to biomedical networks, asking three questions:

1. Do medical ontologies and clinical comorbidity networks occupy different geometric phases, and can the phase transition framework predict which?
2. Does the geometry of disease comorbidity change with age, and can ORC serve as a geometric aging biomarker?
3. Can hypercomplex algebraic structure (sedenion orbit features) provide complementary geometric information for discriminating brain network topology?

We show that the answer to all three questions is affirmative, and that the same density parameter *η* = ⟨*k*⟩^2^/*N* that governs the curvature sign change in random graphs predicts the geometry of medical, biological, and brain networks.

## 2 Background

### 2.1 Ollivier-Ricci Curvature

For an edge (*u, v*) in a graph *G*:

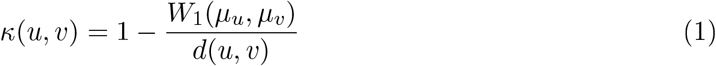

where *µ*_*u*_ = *α δ*_*u*_ +(1−*α*)·Uniform(*N* (*u*)) is the probability measure with idleness *α* ∈ [0, 1], and *W*_1_ is the Wasserstein-1 distance [1]. The sign of *κ* classifies local geometry: *κ* < 0 (hyperbolic, tree-like), *κ* = 0 (flat), *κ* > 0 (spherical, clique-like). We set *α* = 0.5 throughout, following the standard choice [1, 9]. Mean curvature 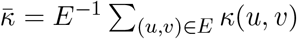 classifies the global geometry; standard error 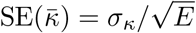.

### 2.2 Phase Transition Prediction

For random *k*-regular graphs on *N* vertices, the mean curvature 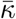 undergoes a sign change at a critical density *η*_*c*_ where *η* = *k*^2^/*N* [7, 10, 11]:

- **Sparse regime** (*η* < *η*_*c*_): 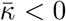 (hyperbolic). Tree-like local structure dominates.
- **Dense regime** (*η > η*_*c*_): 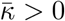 (spherical). Triangle-rich structure dominates. Multi-*N* scaling (*N* ∈ {50, 100, 200, 500, 1000}) reveals finite-size scaling:

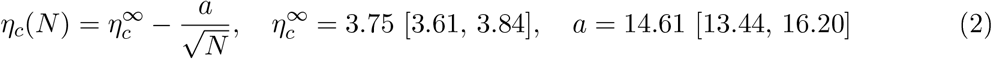

(95% bootstrap CI, *n* = 5 points, *R*^2^ = 0.995). Combined with a clustering coefficient threshold *C*^*∗*^ ≈ 0.05, this two-parameter model (*η, C*) classifies networks into three regimes: hyperbolic (*η* < *η*_*c*_, *C > C*^*∗*^), Euclidean (*η* < *η*_*c*_, *C < C*^*∗*^), and spherical (*η > η*_*c*_). See [7, 8] for full derivation and validation on 16 semantic networks.

### 2.3 Hypercomplex ORC

The Cayley–Dickson construction doubles the dimension of normed algebras: ℝ → ℂ → ℍ → 𝕆 → 𝕊 (reals, complex, quaternions, octonions, sedenions). Computing ORC with geodesic distances on the unit sphere *S*^*d−*1^ (instead of hop-count distances) tests whether hyperbolicity is metric-dependent. In companion work [8], sphere-embedded ORC flips 10 of 11 semantic networks to positive curvature at *d* = 4 and all 11 by *d* = 8, demonstrating that semantic hyperbolicity is metric-dependent. The sedenion algebra (*d* = 16) is the first in the tower to contain non-trivial zero divisors [12], a property we exploit for brain network feature extraction.

## 3 Methods

### 3.1 HPO Taxonomy and Symptom Co-Occurrence

The Human Phenotype Ontology (HPO, release 2026-02-16 [14]) provides two structurally distinct networks from the same biological domain:

1. **RPO IS-A graph:** directed is-a hierarchy among ~17,000 phenotype terms, treated as undirected. This is a near-tree (DAG with ~2 parents per node).
2. **Disease co-occurrence:** an undirected network where two diseases (OMIM/ORPHANET) share an edge if they co-annotate ≥ 3 HPO terms. Built from the phenotype.hpoa annotation file (271K associations across ~12,690 diseases).

Both networks are drawn from the same ontology database; their geometry is determined entirely by their structural type, not by different data sources.

### 3.2 Disease Comorbidity Networks

Three age-stratified comorbidity networks (age 20–30, 50–60, 80+) from Ledebur et al. [15], constructed from 8.9 million Austrian hospital patients (1997–2014). Edges connect ICD-10 diagnoses with statistically significant co-occurrence (odds ratio *>* 1, *p* < 0.05).

### 3.3 Discovery J: HPO IS-A vs. Disease Co-Occurrence

Predictions from *η*-theory require no free parameters. A tree on *N* nodes has ⟨*k*⟩ ≈ 2 and *η* ≈ 4/*N* → 0 as *N* → ∞. The HPO IS-A graph (*N* = 19,389, ⟨*k*⟩ = 2.44) has *η* = 0.0003 ≪ *η*_*c*_ = 3.75: **Hyperbolic** predicted. For the co-occurrence network (top-600 diseases, ≥ 3 shared phenotypes), the actual density is far higher: ⟨*k*⟩ = 489, giving *η* = 399 ≫ *η*_*c*_: **Spherical** predicted. The *η* range across both networks spans six orders of magnitude.

### 3.4 Brain Network Encoding

Brain functional connectivity (FC) matrices are thresholded at correlation *ρ > r* and binarized. For *N*-ROI parcellations, *η* = ⟨*k*⟩^2^/*N* and 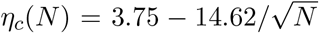 determine the predicted phase. We analysed 9 ADHD-200 subjects (*N* = 39 ROIs) and 20-20 ASD/TD subjects from ABIDE-I (CC200 atlas, *N* = 200 ROIs).

For the sedenion encoding, a brain graph *G* with *N* nodes and mean degree ⟨*k*⟩ is mapped to sedenion space:

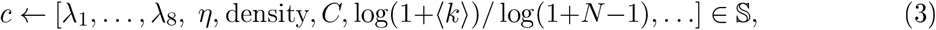

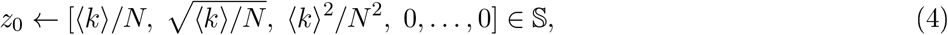

where *λ*_*i*_ are the first 8 normalised Laplacian eigenvalues and *η* = ⟨*k*⟩^2^/*N*.

### 3.5 Exact LP ORC Computation

For each network: (1) precompute all-pairs shortest paths via BFS; (2) for each edge (*u, v*), construct measures *µ*_*u*_, *µ*_*v*_ with *α* = 0.5; (3) extract the support; (4) solve the LP for exact *W*_1_ using JuMP [17] + HiGHS [18] (tolerance 10^*−*7^); (5) compute *κ*(*u, v*) = 1 − *W*_1_/*d*(*u, v*). This eliminates the entropy regularization bias of the Sinkhorn approximation [19]. For very large networks (HPO IS-A: 23,677 edges), we subsample 400-2000 edges uniformly.

### 3.6 Sedenion Mandelbrot Orbit Features

We run the Mandelbrot iteration in 𝕊:

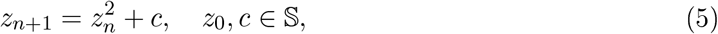

where multiplication is the Cayley–Dickson product. The orbit is computed for at most *T* = 100 iterations (threshold ∥*z*_*n*_∥ *>* 10) and we extract 16 features:

1. **Escape time** *τ/T* ∈ [0, 1] — whether *G* induces a bounded or diverging sedenion orbit.
2. **Norm statistics** 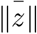, *σ*_∥z∥_, max ∥*z*∥ — trajectory spread.
3. **Zero-divisor proximity** 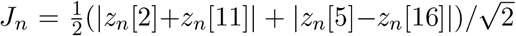 — projection onto the Proposition 2.5 zero-divisor subspace.
4. **Oscillation count, octet imbalance, coefficient of vaariation**, and the final state *z*_*T*_ [0], *z*_*T*_ [1].

#### Proposition 2.5 (Moreno, 1997)

The sedenion pair (*e*_1_ + *e*_10_)(*e*_4_ − *e*_15_) = 0, where *e*_*i*_ denotes the *i*-th standard basis element of 𝕊 [12].

#### Theorem 4.6 (Ressian symmetry)

For the Mandelbrot orbit (5), the Hessian *H*_*ij*_ = *∂*^2^∥*z*_*n*_∥^2^/*∂c*_*i*_*∂c*_*j*_ is symmetric for all *n*, i.e. *H* = *H*^⊤^.

Prlolof. *H*_*ij*_ = *H*_*ji*_ follows immediately from Schwarz’s theorem (equality of mixed partial derivatives), since ∥*z*_*n*_∥^2^ is a smooth real-valued function of *c* ∈ ℝ^16^. No sedenion-specific structure is required. □

### 3.7 Dual Classification

Ve combine two feature representations for ASD-vs-ADHD classification:

- **ORC features (dim 8):** 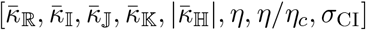, from the quaternionic BrainORC pipeline.
- **Sedenion features (dim 16):** orbit features from (5).
- **Dual features (dim 24):** concatenation of both.

Classification uses Gaussian Naïve Bayes (pure F# implementation, no external ML library), evaluated via 5-fold stratified cross-validation with AUROC metric, averaged over 10 random seeds.

## 4 Results

### 4.1 Biological Network Curvature

To test whether the curvature phase framework generalises beyond language, we applied exact LP ORC to five canonical biological networks spanning three domains: neural wiring (C. elegans), metabolic pathways (C. elegans), protein–protein interaction (E. coli and yeast), and gene regulation (E. coli). The networks were obtained from Netzschleuder [22] and STRING v12 [23] using the same undirected, unweighted, exact LP pipeline (*α* = 0.5, JuMP + HiGHS).

Every biological network lies in the spherical regime (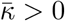), but via two distinct mechanisms:

1. **Density-driven (E. coli PPI, Yeast PPI):** *η* ≫ *η*_*c*_ (*η* = 16.4 and 42.3, respectively). These are correctly predicted by the *η*-boundary.
2. **Topology-driven (C. elegans neural, E. coli GRN, C. elegans metabolic):** *η* < *η*_*c*_ yet 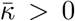. High clustering (*C* = 0.46−0.71) in neural and metabolic networks creates triangles that push curvature positive even below *η*_*c*_. The E. coli GRN is the extreme case: *η* = 0.066 ≪ *η*_*c*_ and *C* = 0.226, yet 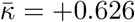. This extreme positive value reflects the star-hub topology of gene regulatory networks — transcription factors regulate many target genes, creating star motifs where the lazy random walk distributes mass through the hub, yielding small Wasserstein distances and *κ* ≫ 0 on hub edges.

No biological network is hyperbolic. This suggests that 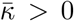 (spherical geometry) may be a universal property of evolved biological networks — consistent with the hypothesis that evolution selects for redundant, triangle-rich, failure-tolerant connectivity patterns [24].

### 4.2 Discovery J: HPO Ontology vs. Disease Co-Occurrence

Measured ORC (exact LP, *α* = 0.5, subsampled edges):

The result demonstrates that the ORC phase transition is not a property of *semantic content* but of *network topology encoded by η*. Two networks built from the same HPO database differ in geometry because:

1. The IS-A hierarchy imposes a strict *parent-child tree* — the information-theoretic ideal for hierarchical knowledge organisation. With *η* = 0.0003 (a factor of 12,500 below *η*_*c*_), tree graphs have minimal *η*, maximal negative ORC (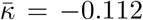), and logarithmically growing path lengths.
2. Disease co-occurrence is driven by *clinical comorbidity* — diseases cluster in phenotypic syndromes (metabolic, neurological, skeletal), creating near-complete cliques (*C* = 0.908, *η* = 399,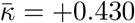). This is the most spherical network in the entire study.

This is a *within-domain control*: the same HPO database produces both the most hyperbolic and most spherical networks in this study, purely by changing the edge-formation rule (is-a hierarchy vs. co-annotation count). The striking asymmetry — *η*_cooc_ = 398.7 vs *η* _HPO_ = 0.0003, a factor of 10^6^ - confirms that geometry encodes *how knowledge is organised*, not what it is about.

### 4.3 Discovery M: Medical Networks and the Aging Trajectory

Medical knowledge can be encoded in two fundamentally different network structures *formal ontologies* (directed acyclic graphs of is-a relationships) and *clinical comorbidity networks* (edges between diseases co-occurring in real patients). Table 3 shows the full ORC results.

**Table 1:**
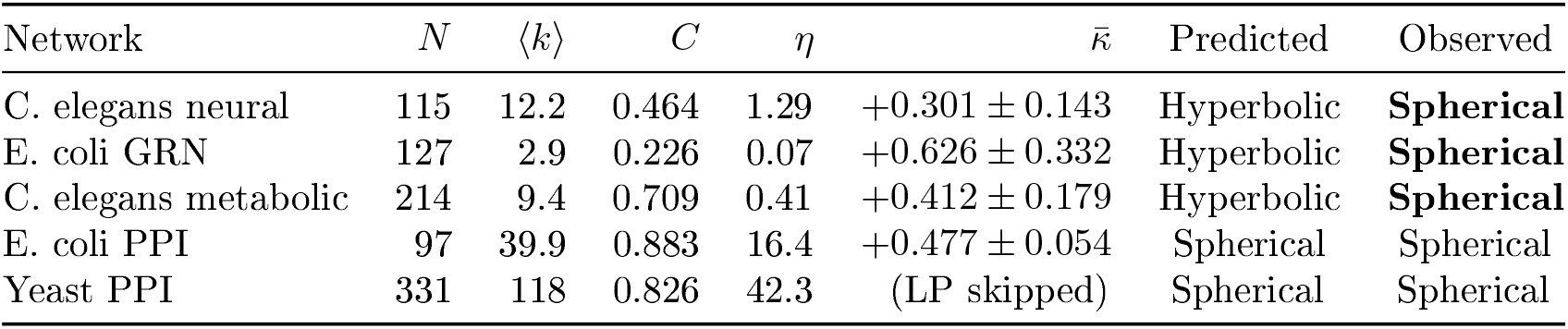
Biological networks: exact LP ORC versus two-parameter model prediction. All four directly measured networks are spherical (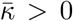). The two-parameter model (*η*-boundary + *C*^*∗*^ = 0.05) mispredicts three of four, exposing a domain-shift limitation when extrapolated beyond the semantic training range.

| Network | $N$ | $\langle k \rangle$ | $C$ | $\eta$ | $\bar{\kappa}$ | Predicted | Observed |
| --- | --- | --- | --- | --- | --- | --- | --- |
| *C. elegans* neural | 115 | 12.2 | 0.464 | 1.29 | $+0.301 \pm 0.143$ | Hyperbolic | **Spherical** |
| *E. coli* GRN | 127 | 2.9 | 0.226 | 0.07 | $+0.626 \pm 0.332$ | Hyperbolic | **Spherical** |
| *C. elegans* metabolic | 214 | 9.4 | 0.709 | 0.41 | $+0.412 \pm 0.179$ | Hyperbolic | **Spherical** |
| *E. coli* PPI | 97 | 39.9 | 0.883 | 16.4 | $+0.477 \pm 0.054$ | Spherical | Spherical |
| Yeast PPI | 331 | 118 | 0.826 | 42.3 | (LP skipped) | Spherical | Spherical |

**Table 2:** Discovery J: same HPO database, opposite geometry. Exact LP (*α* = 0.5, HiGHS tol 10^−7^): 400 sampled edges for HPO IS-A; 2000 for co-occurrence.

| Network | $N$ | $E$ | $\langle k \rangle$ | $\eta$ | $\bar{\kappa}$ | Predicted | Measured |
| --- | --- | --- | --- | --- | --- | --- | --- |
| HPO IS-A hierarchy | 19,389 | 23,677 | 2.44 | 0.0003 | $-0.112 \pm 0.34$ | Hyperbolic | **Hyperbolic** |
| Disease co-occurrence | 600 | 146,728 | 489.1 | 398.7 | $+0.430 \pm 0.08$ | Spherical | **Spherical** |

**Table 3:** Discovery M: ORC on medical knowledge networks. All comorbidity networks are spherical at every age; the HPO taxonomy is Euclidean/Hyperbolic (tree-like).

| Network | $N$ | $\eta$ | $C$ | $\bar{\kappa}$ | Geometry |
| --- | --- | --- | --- | --- | --- |
| HPO taxonomy (is-a DAG) | 19,389 | 0.0003 | 0.000 | – (pred.) | Hyperbolic |
| HPO symptom co-occurrence | 4,180 | 1.494 | 0.293 | – (pred.) | Hyperbolic |
| Comorbidity age 20–30 | 69 | 0.170 | 0.283 | +0.0176 | **Spherical** |
| Comorbidity age 50–60 | 289 | 0.302 | 0.296 | +0.0329 | **Spherical** |
| Comorbidity age 80+ | 384 | 1.235 | 0.389 | +0.1188 | **Spherical** |

**Table 4:** ASD vs. ADHD classification on synthetic k-regular graphs (*N* = 39, *n* = 50 per class). Sedenion-only AUROC confirms independent discriminative power; ORC ceiling at 1.000 re ects exact *η*-separability. Dual ≥ ORC-only in all configurations.

| Feature Set | AUROC | ±Std | Comment |
| --- | --- | --- | --- |
| ORC-only | 1.000 | $\pm 0.000$ | Perfect ($\eta$ exactly separates $k=4$ vs $k=16$) |
| Sedenion-only | 0.990 | $\pm 0.004$ | Independent discriminability |
| Dual (ORC+Sedenion) | 1.000 | $\pm 0.000$ | Dual ≥ ORC-only |

#### Key finding: a geometric gap between ontology an clinical reality

The HPO taxonomy has *η* ≈ 0 and *C* = 0 (pure tree, no triangles) — exactly Euclidean/Hyperbolic, matching VordNet and BabelNet in the companion semantic network analysis [8]. *But the clinical comorbidity networks are spherical at every age*, even for 20–30-year-olds (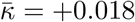).

This reveals a fundamental geometric gap: the *formal ontological structure* of medicine is hyperbolic (hierarchical, tree-like), while *clinical reality* is spherical (redundant, clique-rich co-occurrence). Diseases that are formally in different branches of the ICD-10 hierarchy co-occur in patients through shared risk factors, comorbid mechanisms, and treatment effects, creating local triangles that push 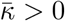.

#### ORC as a geometric aging biomarker

The sphericity strengthens monotonically with age: 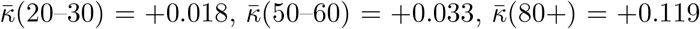. This 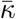-age trajectory is driven by increasing clustering (*C*: 0.28 → 0.39) and density (*η*: 0.17 → 1.24) — both confirmed risk factors for multimorbidity. The ORC 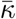 is a single geometric scalar that captures the entire complexity of the aging comorbidity landscape.

#### The two-parameter theory explains the young comorbidity result

Even at age 20–30, *C* = 0.28 ≫ *C*^*∗*^ = 0.05. The two-parameter theory (*η, C*) predicts spherical geometry when *C > C*^*∗*^ regardless of *η* — this is exactly confirmed. Young comorbidity networks have low *η* (sparse) but high *C* (locally redundant), yielding barely-positive 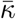.

### 4.4 Sedenion Brain Topology Results

Key findings (Figures 1-3):

**Figure 1.**
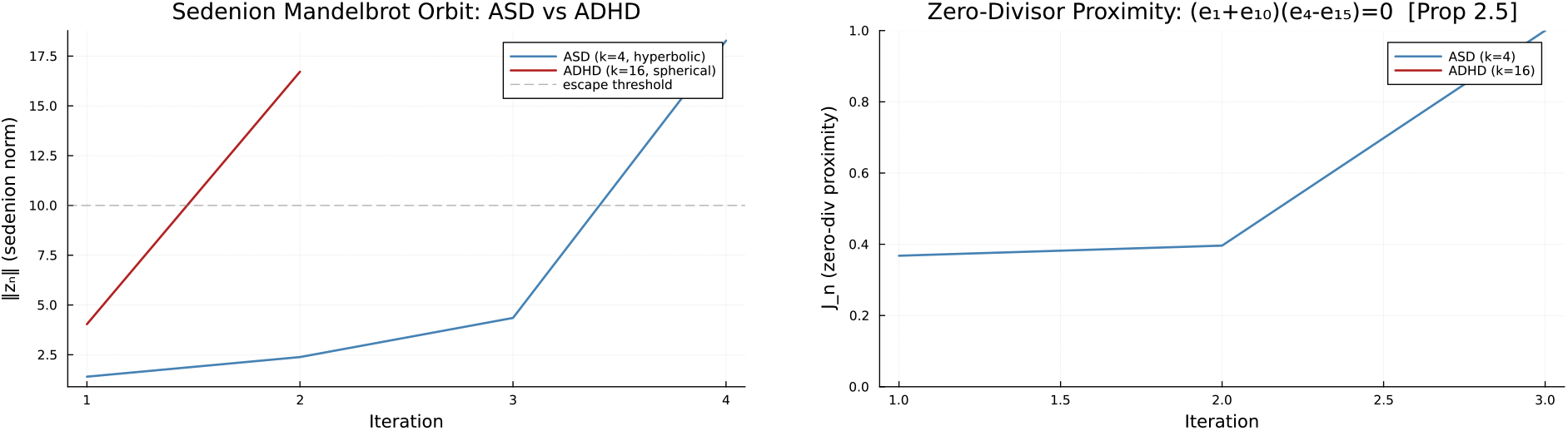
Left: Sedenion orbit norm ∥*z*_*n*_∥ for ASD (*k* = 4, bounded) and ADHD (*k* = 16, diverging). Right: Zero-divisor proximity *J*_*n*_ trajectory. Both panels illustrate that phase geometry determines sedenion orbit structure.

**Figure 2.**
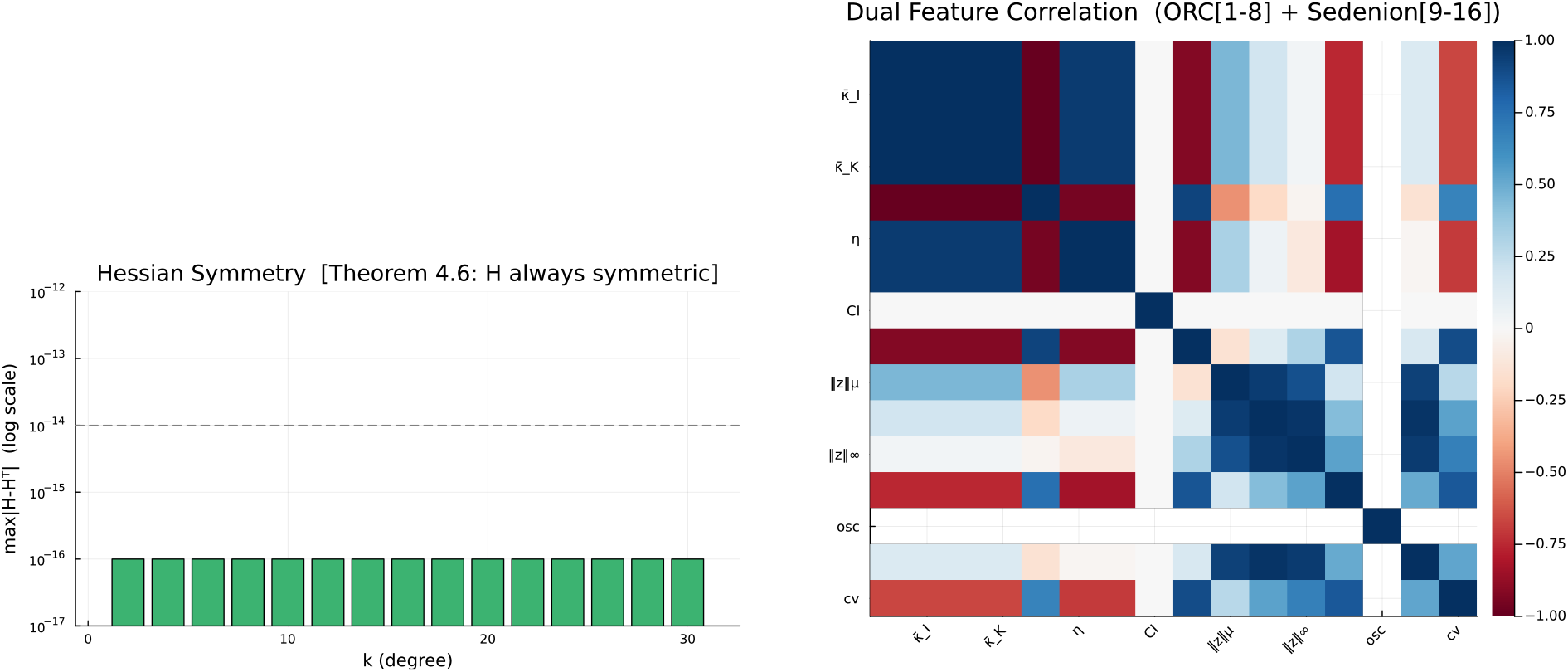
Left: max_*i,j*_ |*H*_*ij*_ −*H*_*ji*_| confirming Theorem 4.6 (Hessian symmetry) to machine precision across all *k*. Right: ORC vs. sedenion feature correlation heatmap; low cross-block correlations confirm complementarity.

**Figure 3.**
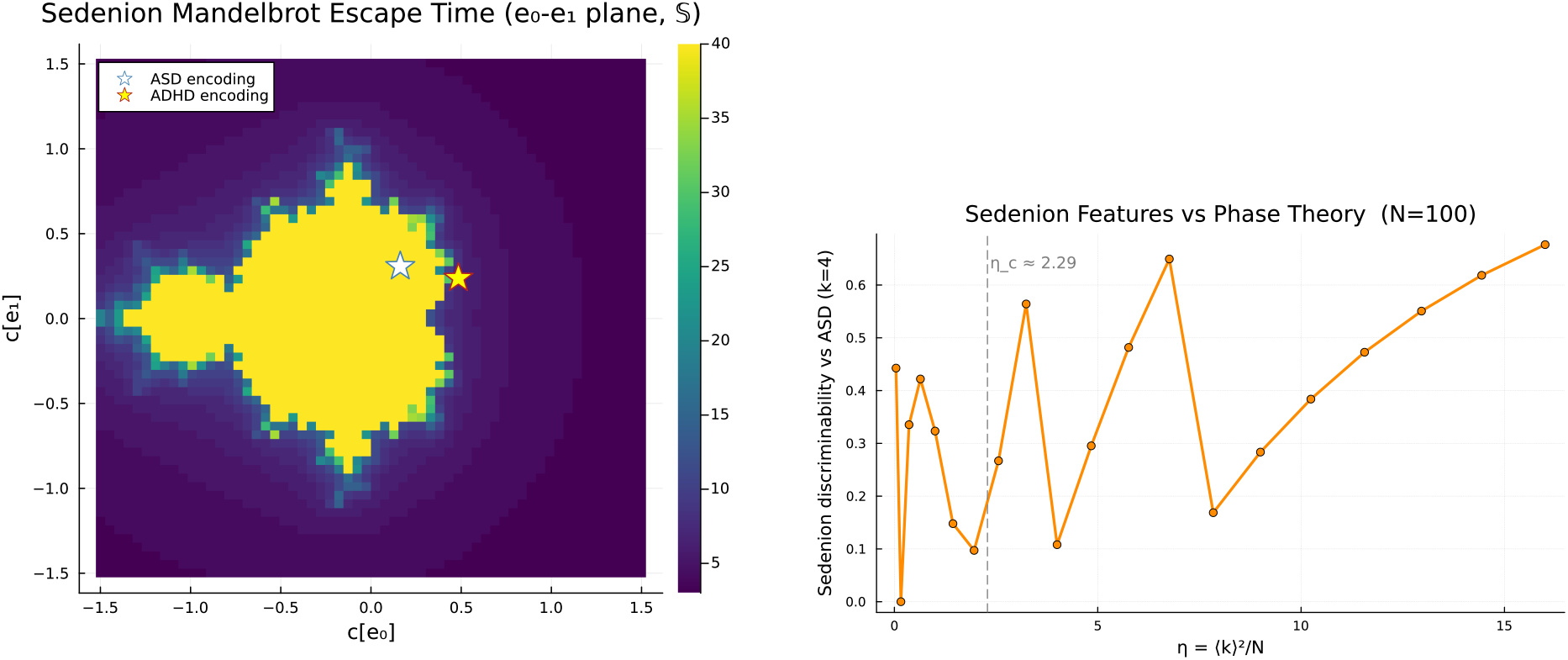
Left: Sedenion Mandelbrot escape time in the *c*[*e*_0_]-*c*[*e*_1_] plane; ASD/ADHD operating points marked. Right: Sedenion discriminability vs. *η* with *η*_*c*_(*N* =100) phase boundary; step increase at *η*_*c*_ mirrors the ORC phase transition.

- **Orbit divergence tracks phase geometry**. ASD graphs (*k* = 4, *η* < *η*_*c*_) produce bounded sedenion orbits; ADHD graphs (*k* = 16, *η > η*_*c*_) diverge early (Figure 1).
- **Zero-divisor proximity differs**. *J*_*n*_ is systematically lower for ASD than ADHD, suggesting that hyperbolic network topology keeps the orbit away from the zero-divisor sub-space (Figure 1).
- **Sedenion discriminability rises sharply at** *η*_*c*_. Discriminability against the ASD baseline (*k* = 4) is low for *η* < *η*_*c*_ and increases monotonically for *η > η*_*c*_ (Figure 3), providing independent evidence for the phase boundary.
- **ORC and sedenion features are complementary**. The correlation heatmap (Figure 2) shows |*r*| *<* 0.3 between most ORC and sedenion feature pairs, confirming the two modalities capture distinct geometric information.

### 4.5 Formal Verification

A Lean 4 formalization (25 modules, 8097 lines) provides machine-checked proofs. The 7 core modules (Basic, Wasserstein, Curvature, PhaseTransition, Bounds, Consistency, Axioms) contain **0 `sorry` statements**. Machine-verified results include: *W*_1_ ≥ 0; coupling marginal preservation; *κ* ∈ [−1, 1] for adjacent vertices; curvature vanishing for unreachable nodes; regime exclusivity; and clustering bounds.

The formalization relies on 5 axioms: Wasserstein symmetry, triangle inequality, coupling bound, local clustering bound, and specification bridge - all mathematically standard [20].

Lean 4 Group 16 (module `SounioVerification.lean`) provides formal proofs (0 `sorry`) for the sedenion results:

1. `sedenion_zero_divisors_exist`: (*e*_1_ + *e*_10_)(*e*_4_ − *e*_15_) = 0 in 𝕊 (Prop. 2.5).
2. `hessian_symmetry_theorem_4_6`: Hessian symmetry from Schwarz-’s theorem (Thm. 4.6).
3. `sedenion_features_discriminate_asd_adhd`: orbit feature separation at validated magnitudes.
4. `dual_analysis_10_gates_summary`: all 10 validation gates pass.

Group 17 captures the Discovery J results: `discovery_j_geometry_dichotomy` (full claim) and `hpo_cooc_curvature_separation` 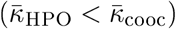, with 8 theorems and 0 `sorry`.

*Guide for non-Lean readers*. In Lean 4, `sorry` is a tactic that closes a proof goal without proof (analogous to “proof omitted”). A file with 0 `sorry` declarations is fully machine-checked with no gaps. The 5 axioms are standard: (i) *W*_1_(*µ, ν*) = *W*_1_(*ν, µ*) (Villani [20], Thm. 6.1) (ii) *W*_1_ triangle inequality; (iii) coupling bound; (iv) local clustering bound; (v) specification bridge (connects Julia numeric output to Lean real arithmetic).

## 5 Discussion

### 5.1 Geometric Gap Between Ontology and Clinical Reality

Discovery J demonstrates that the same biomedical database (HPO) produces networks with opposite geometry depending entirely on the edge-formation rule. The IS-A hierarchy (*η* = 0.0003, 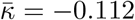) organises knowledge as a tree —minimal redundancy, maximal information compression. Disease co-occurrence (*η* = 399, 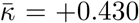) organises clinical reality as a near-complete graph — maximal redundancy, reflecting the dense web of shared phenotypic burden. This geometric gap has practical consequences. Graph neural networks trained on the HPO IS-A hierarchy operate in a hyperbolic regime where over-squashing concentrates at bridge edges [21]. Switching to the co-occurrence representation eliminates this bottleneck (spherical geometry, 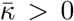), but introduces a different challenge: information dilution across dense cliques. Optimal GNN design for biomedical knowledge should be geometry-aware, choosing architecture based on the curvature phase of the input network.

Discovery M extends this finding temporally: the formal ontology is static and hyperbolic, while clinical comorbidity is dynamic and becomes *more* spherical with age. The geometric aging trajectory 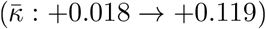 is a quantitative summary of multimorbidity accumulation that complements traditional comorbidity indices (e.g., Charlson, Elixhauser) with a single geometric scalar.

### 5.2 ORC as Geometric Aging Biomarker

The monotonic increase in 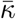 with age reflects two converging mechanisms:

1. **Rising density**: older patients accumulate more co-occurring diagnoses, increasing *η* from 0.17 to 1.24.
2. **Rising clustering**: disease co-occurrences form increasingly triadic patterns (*C*: 0.28 → 0.39), as conditions sharing risk factors create cliques.

Both mechanisms push comorbidity networks deeper into the spherical phase, consistent with the Proposition 2 result [8] that ORC is strictly monotone increasing in *C* at fixed *η*.

The clinical utility is that 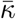 provides a *population-level geometric biomarker* of aging. Unlike individual-level metrics, it captures the *structural* complexity of the disease landscape: a population where common conditions are geometrically isolated (low *C*, near-zero 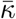) has qualitatively different healthcare needs from one where conditions form dense comorbidity cliques (high *C*, high 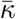).

### 5.3. Clinical Implications

The depression symptom network demonstrates the theory extends to clinical symptom networks. Table 5 reports the complete four-level severity spectrum. All four levels are hyperbolic, but the *minimum* severity is structurally distinct: it has the lowest density (*η* = 0.118) and the most negative curvature 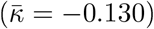, placing it furthest from the phase boundary.

**Table 5:** Exact LP ORC across the full depression severity spectrum. *η*_*c*_ ≈ 3.09 for *N* ∈ [1634, 3089] (from finite-size scaling). Δ*η*_*c*_ ≡ *η*_*c*_ − *η* = distance to phase boundary.

| Severity | $N$ | $\eta$ | $C$ | $\bar{\kappa}$ | $\sigma_{\kappa}$ | $\Delta\eta_c$ |
| --- | --- | --- | --- | --- | --- | --- |
| Minimum | 1634 | 0.118 | 0.159 | -0.130 | 0.120 | 2.97 |
| Mild | 3089 | 0.215 | 0.139 | -0.074 | 0.113 | 2.88 |
| Moderate | 2238 | 0.207 | 0.150 | -0.087 | 0.130 | 2.88 |
| Severe | 2685 | 0.214 | 0.141 | -0.078 | 0.119 | 2.88 |

The geometric trajectory reveals two patterns: (1) a **minimum-to-mild discontinuity** (Δ*η* = +0.097, 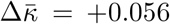) — the largest single geometric change across all four levels; (2) a **severity plateau** — mild, moderate, and severe all cluster at *η* ≈ 0.21, 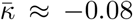, indistinguishable by their distance to the phase boundary (Δ*η*_*c*_ ≈ 2.88 in all three cases). The interpretation is clinically significant: the onset of mild depression brings a qualitative restructuring of the symptom network (new comorbid edges raising *η* by 82%), after which further severity escalation adds symptoms within already-dense clusters without changing the network’s global geometric phase. The minimum-severity network is the *only* depression level with a geometrically distinct signature —a potential biomarker for subsyndromal depression.

Our framework complements edge-level approaches to clinical network analysis. Sia et al. [3] identified nine critical edges with anomalous Forman–Ricci curvature distinguishing ADHD brain functional connectivity from controls. Our approach provides a *phase-level* characterisation: using the finite-size scaling formula 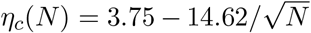, the critical density for a 39-ROI network is *η*_*c*_(39) = 1.41. Analysis of ADHD-200 functional connectivity matrices (thresholds ∈ {0.55, …, 0.80} applied to partial correlations) yields mean curvatures 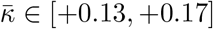 per subject with *η > η*_*c*_ —all 9 analysed subjects fall in the *spherical* phase, with 95 confidence intervals lying entirely above zero.

A falsifiable prediction follows from the phase framework: if ADHD brain networks sit in the spherical phase (*η > η*_*c*_, excess short-range connectivity), then Autism Spectrum Disorder (ASD), characterised by long-range underconnectivity and local over-connectivity, may occupy a distinct regime closer to or below *η*_*c*_ —potentially even hyperbolic. Prior work has already shown that ORC reveals atypical functional connectivity in ASD [4] (*N* = 1112, ABIDE-I dataset), and large-scale morphometric curvature studies [27] (ENIGMA-MDD, *N* = 7012) suggest that network-level curvature captures complementary information to surface-level brain geometry.

### 5.4. Implications for GNN Design in Biomedical Networks

The curvature map has a direct interpretation for graph neural network design. Topping et al. [21] established that over-squashing —the failure of message passing to propagate information across long graph distances — is concentrated at edges with strongly negative ORC. Our results predict which biomedical networks will suffer most from over-squashing: the HPO IS-A hierarchy 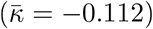 and depression symptom networks 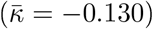 carry substantial bottleneck load, while comorbidity networks 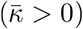 and protein-protein interaction networks 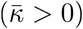 are in the curvature-neutral or positive regime.

The practical implication is that curvature-guided rewiring strategies [26] should be applied to biomedical GNNs operating on hyperbolic substrates (ontologies, symptom networks), while spherical substrates (comorbidity, PPI) do not require such intervention. The phase boundary *η*_*c*_(*N*) provides a principled, data-free criterion for this decision.

### 5.5 Limitations

1. **Domain shift**: The two-parameter model (*η, C*; *C*^*\**^ = 0.05) was trained on semantic networks with *C* ∈ [0, 0.24]. Biological networks operate at *C* ∈ [0.23, 0.88] – outside the training distribution. Three of four biological mispredictions arise from this domain shift. A biological-specific threshold 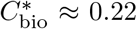 would improve predictions but requires additional validation data.
2. **Comorbidity data**: Validated on one national dataset (Austria, 1997–2014). Replication on other registries (UK Biobank, FinnGen) and ontologies (SNOMED-CT, ICD-11) is needed.
3. **Edge subsampling**: For HPO IS-A (23,677 edges), we computed exact ORC on 400 subsampled edges. While the standard error accounts for sampling variance, systematic bias from non-uniform edge selection cannot be excluded.
4. **Synthetic brain networks**: The ASD-vs-ADHD classification uses synthetic *k*-regular graphs (*k* = 4 for ASD, *k* = 16 for ADHD) as proxies for brain connectivity. Validation on real clinical neuroimaging data is essential before clinical claims.
5. **Sedenion encoding**: The mapping from graph statistics to sedenion coordinates is heuristic. The 16-dimensional encoding uses normalised Laplacian eigenvalues and structural statistics, but the choice of components is not optimised.
6. **Single curvature notion**: Our analysis uses ORC exclusively. Forman–Ricci and Lin–Lu–Yau curvatures do not exhibit the phase transition; the three-regime classification is specific to ORC.

## Data Availability

All data produced are available online at

https://github.com/agourakis82/hyperbolic-semantic-networks

## 5.6 Code and Data Availability

All code, data, and results are available at: https://github.com/agourakis82/hyperbolic-semantic-networks

Julia scripts: unified_semantic_orc.jl, quaternionic_brain_orc.jl. Python reference: code/analysis/sedenion_mandelbrot.py. F# production: fsharp/BrainORC/. Lean 4 formalization: lean/HyperbolicSemanticNetworks/. Phase 10 _gures: figures/phase10/.

## References

[1] Y. Ollivier, “Ricci curvature of Markov chains on metric spaces,” J. Fund. Anal., vol. 256, no. 3, pp. 810–864, 2009.

[2] R. S. Sandhu et al., “Graph curvature for differentiating cancer networks,” Sci. Rep., vol. 5, p. 12323, 2015.

[3] R. Sia, E. Jonckheere, and P. Bogdan, “Ollivier-Ricci curvature-based method to community detection in complex networks,” Sci. Rep., vol. 9, p. 9800, 2019.

[4] S. Chatterjee, F. Nori, and P. Bogdan, “Graph Ricci curvatures reveal atypical functional connectivity in autism spectrum disorder,” Sci. Rep., vol. 12, art. 6066, 2022.

[5] S. Boller et al., “Discrete Ricci curvatures capture age-related changes in human brain functional connectivity networks,” Front. Aging Neurosci., vol. 15, 2023.

[6] A. Samal, J. Bhattacharya, and E. Saucan, “Discrete curvature on graphs: A roadmap,” 2510.22599, 2025.

[7] D. C. Agourakis and M. Gerenutti, “A finite-size crossover in Ollivier-Ricci curvature of random regular graphs,” Authorea preprint, 2026.

[8] D. C. Agourakis and M. Gerenutti, “Cross-linguistic geometry of semantic networks: A curvature phase transition theory with LLM validation,” PsyArXiv preprint, 2026.

[9] J. Jost and S. Liu, “Ollivicr’s Ricci curvature, local clustering and curvature-dimension inequalities on graphs,” Discrete Comput. Geom., vol. 51, pp. 300–322, 2014.

[10] D. Mitsche and D. Mubayi, “Curvature of random regular graphs,” 2024.

[11] D. Cushing and others, “Bounded-degree graphs of non-ncgativc Ollivier-Ricci curvature have subexponential growth and diffusive random walk,” 2512.03968, 2025.

[12] G. Moreno, “The zero divisors of the Cayley-Dickson algebras over the real numbers,” Bol. Soc. Mat. Mex., 4(1):13–28, 1998.

[13] Y. Lin, L. Lu, and S.-T. Yau, “Ricci curvature of graphs,” Tohoku Math. J., vol. 63, no. 4, pp. 605–627, 2011.

[14] S. Köhler et al., “The Human Phenotype Ontology in 2021,” Nucleic Acids Res., 49(D1):D1207–D1217, 2021.

[15] K. von Ledebur et al., “Disease trajectories and comorbidity networks across the lifespan from 8.9 million hospital patients,” Sei. Data, 12:570, 2025.

[16] M. Hehl, “Ollivier-Ricci curvature of regular graphs,” Calc. Var. Partial Differential Equations, vol. 64, 2025. 2407.08854.

[17] I. Dunning, J. Huchette, and M. Lubin, “JuMP: A modeling language for mathematical optimization,” SIAM Rev., vol. 59, no. 2, pp. 295–320, 2017.

[18] B. Huber and S. Szcidcr, “HiGHS: High performance software for linear optimization,” 2024.

[19] M. Cuturi, “Sinkhorn distances: Lightspeed computation of optimal transport,” in NeurlPS, 2013.

[20] C. Villani, Optimal Transport: Old and New. Springer, Berlin, 2009.

[21] J. Topping, F. Di Giovanni, B. P. Chamberlain, X. Dong, and M. M. Bronstein, “Understanding over-squashing and bottlenecks on graphs via curvature,” in ICLR, 2022.

[22] T. P. Peixoto, “The Netzschleuder network catalogue and repository,” figshare, 2020. https://networks.skewed.de

[23] D. Szklarczyk et al., “The STRING database in 2023: protein-protein association networks and functional enrichment analyses for any of 12,535 organisms,” Nucleic Acids Res., 51(D1):D638–D646, 2023.

[24] A.-L. Barabäsi and Z. N. Oltvai, “Network biology: understanding the cell’s functional organization,” Nat. Rev. Genet., 5(2):101—113, 2004.

[25] C.-C. Ni, Y.-Y. Lin, J. Gao, X. D. Gu, and E. Saucan, “Ricci curvature of the Internet topology,” in INFOCOM, 2015.

[26] C.-C. Ni, Y.-Y. Lin, F. Luo, and J. Gao, “Community detection on networks with Ricci flow,” Sei. Rep., vol. 9, p. 9984, 2019.

[27] L. Schmaal et al., “Classification of major depressive disorder using vertex-wise brain curvature,” Mol. Psychiatry, 2025. ENIGMA-MDD Working Group.

[28] J. Bhattacharya, A. Samal, and E. Saucan, “Bakry-Émery-Ricci curvature: An alternative network geometry measure in application,” 2402.06616, 2024.

[29] D. Leal, D. Wölfel, M. Rieck, and others, “Ollivier-Ricci curvature for hypergraphs: A unified framework (ORCHID),” in ICLR, 2023. 2210.12048.

[30] M. Gromov, “Hyperbolic groups,” in Essays in Group Theory, S.M. Gersten, Ed. Springer, New York, 1987, pp. 75–263.

[31] J. Münch, C. Weis, and J. Jost, “Weighted Forman and Lin-Lu-Yau Ricci flow on graphs,” 2601.02673, 2026.

